# Reduced dementia incidence following varicella zoster vaccination in Wales 2013–2020

**DOI:** 10.1101/2021.07.22.21260981

**Authors:** Christian Schnier, Janet Janbek, Richard Lathe, Jürgen Haas

## Abstract

**INTRODUCTION:** Chronic infection with herpes viruses is a potential contributing factor to the development of dementia. The introduction of nationwide shingles (varicella zoster) vaccination in Wales might therefore be associated with reduced incident dementia.

**METHODS:** We analyzed the association of shingles vaccination with incident dementia in Wales between 2013 and 2020 using retrospectively collected national health data.

**RESULTS:** Vaccinated individuals were at reduced risk of dementia (adjusted hazard ratio: 0.72; 95% CI: 0.69 to 0.75). The association was not modified by a reduction in shingles diagnosis and was stronger for vascular dementia than for Alzheimer’s disease. Vaccination was also associated with a reduction in several other diseases and all-cause mortality.

**DISCUSSION:** Our study shows a clear association of shingles vaccination with reduced dementia, consistent with other observational cohort studies. The association may reflect selection bias with people choosing to be vaccinated having a higher healthy life expectancy.

## 1. Introduction

Research into the association of vaccination with dementia serves several purposes. Any association would highlight the contribution of the immune system and/or of pathogens to the disease progression to dementia and would allow a deeper understanding of the natural history of dementia. In addition, vaccination strategies could be developed to cost-effectively reduce the rate of dementia.

Several observational cohort and case–control studies have shown a reduction in dementia rates post-vaccination. Twenty years ago Verreault *et al*.[1] reported that reported that vaccine exposure (diphtheria/tetanus, polio, influenza) was associated with a 25–60% reduction in later Alzheimer’s disease (AD) development. Klinger *et al*.[2] demonstrated a significantly reduced risk of developing AD in bladder cancer patients exposed to repeated intravesicalar applications of Bacillus Calmette– Guérin (BCG) vaccine, especially in the population aged 75 years and older. Scherrer *et al*. showed a significantly reduced rate of dementia in people vaccinated with Tdap[3] and shingles vaccination compared to those not vaccinated using data from two American disease registers (Veterans Health Affairs and MarketScan). Liu *et al*.[4] found a reduced dementia rate in chronic kidney disease patients vaccinated with influenza vaccine using data from the National Health Insurance Research Database of Taiwan. However, observational studies to ascertain vaccine efficacy are not easy to interpret[5], and, to our knowledge, no vaccine, whether specific for dementia or with a primary target other than dementia, has been proven in a clinical trial to be efficient in preventing dementia.

Chronic infection with human herpes viruses (HHVs) has recently been highlighted as a potential contributing factor to the development of dementia, especially AD[6]. Population-wide observational cohort studies indicate a moderate to non-existing positive association of a diagnosis of HHV infection with incident dementia, and some studies point to a potential mediating role of antiherpetic medication [7–10].

HHV3, also known as varicella zoster virus (VZV), is generally acquired early in childhood when it causes chickenpox, but the virus persists lifelong and can re-emerge in the elderly as shingles, and has also been associated with postherpetic neuralgia, encephalitis and/or meningitis, and respiratory disease[11]. To reduce the effects of HHV3 re-emergence in the elderly, national vaccination strategies have been implemented in the UK and elsewhere. In Wales, national shingles vaccination has been conducted since 2013, with the aim to vaccinate people aged 70 years, and a catch-up vaccination at age 79 for those not vaccinated at age 70[12]. Until 2018 the only available shingles vaccine in Wales was an attenuated live HHV3 vaccine (Zostavax); since June 2018 a small proportion of the Welsh population received recombinant shingles vaccine (Shingrix).

Our objective was to analyze the association of shingles vaccination with incident dementia in Wales between 2013 and 2020. Furthermore, we analyzed whether that association was mediated by a reduction of diagnosed shingles and whether the association was different for AD than for vascular dementia (VaD).

## 2. Material and Methods

### 2.1. Study population and period

The study population comprised the complete population of Wales born after 01.09.1933 who were registered with a primary care provider (general practitioner, GP) and whose GP was reporting into the Secure Anonymised Information Linkage Databank (SAIL) databank in Wales (approximately 80%[13]). For each person in the study population, follow-up started with a 70th birthday at any time during the period between 2013–2020; however, to allow for retrospective classification of risk factors before vaccination, people were excluded who, at their 70th birthday, were not registered with a participating GP for at least 5 years. People were also excluded for whom date of birth, sex, or socioeconomic status was missing or who were diagnosed with dementia before their 70th birthday.

### 2.2. Classification

Vaccination status, dementia incidence, and covariates used in the analyses were classified using routinely collected health data (RCHD) in the SAIL databank[14]. The databank holds linked information from primary care (READ V2), hospital admissions (ICD 10), and mortality records (ICD 10), as well as registration data and information on area-based socioeconomic status (Welsh Index of Multiple Deprivation, WIMD).

Dementia was classified using primary care diagnostic data and hospital admission data, and the first code found was used as the date of diagnosis. The codes used to classify dementia (Table 1 in the supplementary material online) in British RCHD have been validated previously and found to be of good predictive value[15]. Medication codes were not used because we judged that the positive predictive value for a medication code in the absence of a concurrent diagnostic code to be insufficient. Exposure to the shingles vaccine was classified using codes from the primary care records, either as a shingles (herpes zoster) vaccination (preventive procedure) or as a prescription for a shingles vaccination. The validity of the codes for shingles vaccination (Table 1 in the supplement) has to our knowledge not been studied, but the same data source and codes are used for national vaccine coverage reports by the Public Health Wales Vaccine Preventable Disease Programme. The attenuated live herpes zoster vaccine used for most people in the study population is given only once; in the unusual event of a second vaccination (mostly recombinant zoster vaccine) we have taken the time of first vaccination as time of exposure. Shingles and other diseases were classified using primary care diagnostic data and hospital admission data; again, medication codes on their own were deemed to be insufficient. Care home residency between age 65 and 70 years (before follow-up) was classified using codes from the social/personal history recorded in the primary care data; previous research has shown that recording of care home residence is limited in primary care records[16]. Frailty was classified using a summary measure of 5 years of GP data between the ages of 65 and 70, including 36 ’deficits’ (clinical signs, symptoms, diseases, and disabilities)[17]. Finally, we included 11 comorbidities that make up the Charlson comorbidity index, again classified over 5 years of GP data between the ages of 65 and 70. In contrast to the calculation of the electronic frailty index (EFI) as published[17], and the Charlson co-morbidity index as published[18], in our study a diagnosis of dementia was excluded.

**Table 1.** Association of risk factors with exposure to shingles vaccine.

| **Variable** | **Level<sup>†</sup>** | **Total N** | **Vaccinated N (%)** | **Hazard ratio (95% CI)<sup>\*</sup>** |
| --- | --- | --- | --- | --- |
| Sex | Female | 174317 | 81064 (47%) | 0.95 (0.95 to 0.96) |
| Born after 1943 | Yes | 161476 | 90012 (56%) | 26.63 (25.99 to 27.29) |
| WIMD<sup>†</sup> | 1 | 54950 | 22576 (41%) | 1.00 (ref) |
|  | 2 | 65067 | 27855 (43%) | 1.04 (1.02 to 1.06) |
|  | 3 | 70425 | 31499 (45%) | 1.08 (1.07 to 1.1) |
|  | 4 | 67269 | 33150 (49%) | 1.22 (1.2 to 1.24) |
|  | 5 | 78748 | 40947 (52%) | 1.28 (1.26 to 1.3) |
| Frailty | Fit | 255118 | 119192 (47%) | 1.00 (ref) |
|  | Mild | 69379 | 32234 (46%) | 1.08 (1.07 to 1.09) |
|  | Moderate | 10942 | 4291 (39%) | 0.99 (0.96 to 1.02) |
|  | Severe | 1020 | 310 (30%) | 0.85 (0.76 to 0.95) |
| Care home | Yes | 432 | 89 (21%) | 0.57 (0.46 to 0.7) |
| Prior vaccination | Yes | 264454 | 141164 (53%) | 3.01 (2.96 to 3.06) |
| Diabetes | Yes | 47411 | 22232 (47%) | 1.13 (1.11 to 1.14) |
| Cancer | Yes | 23987 | 11085 (46%) | 1.07 (1.05 to 1.1) |
| Cerebrovascular disease | Yes | 10690 | 4228 (40%) | 0.87 (0.84 to 0.9) |
| Chronic obstructive pulmonary disease | Yes | 37902 | 18449 (49%) | 1.13 (1.11 to 1.14) |
| Chronic heart disease | Yes | 4858 | 1815 (37%) | 0.89 (0.85 to 0.94) |
| Chronic liver disease | Yes | 1000 | 370 (37%) | 0.95 (0.85 to 1.05) |
| Myocardial infarction | Yes | 5514 | 2231 (40%) | 0.83 (0.79 to 0.86) |
| Peptic ulcer | Yes | 2085 | 813 (39%) | 0.77 (0.72 to 0.82) |
| Perivascular disease | Yes | 5695 | 2075 (36%) | 0.84 (0.81 to 0.88) |
| Renal disease | Yes | 20694 | 9165 (44%) | 0.95 (0.93 to 0.97) |
| Rheumatic disease | Yes | 6444 | 2411 (37%) | 0.77 (0.74 to 0.8) |

<sup>\*</sup>For sex, birth year, Care home, prior vaccination, and all disease groups, the reference (HR=1) is set to males, born before 1943, not in a care home, no prior vaccination, and people without each of the diseases, respectively.
<sup>†</sup>Welsh Index of Multiple Deprivation: level 1 is most deprived, 5 is least deprived.

### 2.3. Statistical analysis

We first identified factors associated with vaccination and with incident dementia in the scientific literature. We then analyzed the association of multiple putative risk factors with shingles vaccination in several univariable Cox proportional hazard models (CoxPH). We used factors associated with vaccination in addition to other reported risk factors for dementia in several univariable CoxPH models on the hazard of incident dementia. We used fractional polynomials to find the best transformation for the EFI and added it to the model as a continuous variable; all other variables were categorical. Year of birth was categorized as <1943 and later because those born before 1943 were only eligible for the catch-up vaccination. All survival analysis models were stratified by that category to allow for different underlying hazard functions. Finally, we modeled the association of shingles vaccination with incident dementia in a multivariable CoxPH model in which shingles vaccination was included as time-dependent variable. To adjust for correlation between patients of the same GP practice, we added the practice number as a random effect (frailty model). In all survival models, people were followed up from their 70th birthday to the first diagnosis of dementia or censoring (death or the end of the study period in January 2020).

To analyze whether the association was different for AD than for VaD, we replaced the dementia outcome with the more specific outcome of either AD or VaD. In these models we additionally censored observations at the time of any other dementia.

To analyze the association of vaccination with shingles incidence, we used a similar CoxPH model with shingles vaccination included as a time-dependent variable and a shingles diagnosis as the outcome variable. In that model, to avoid effects of an increased shingles risk in people with dementia, we additionally censored observations at the time of dementia.

To analyze if the association of vaccination with dementia was mediated by a reduction in shingles incidence in those vaccinated, we modeled the association of shingles vaccination with incident dementia in a multivariable CoxPH model, with shingles vaccination and shingles diagnosis included as time-dependent variables. This created five different exposure categories: (i) not vaccinated, no shingles diagnosis (every person at the start of follow-up); (ii) not vaccinated, shingles diagnosed; (iii) vaccinated, no shingles diagnosed; (iv) first vaccinated then shingles diagnosed; and (v) first shingles diagnosis then vaccinated.

To test, whether the association of the shingles vaccine was specific for dementia, we additionally analyzed the association of vaccination with cancer, myocardial infarction (MI), stroke, hip fracture, and all-cause mortality. These outcomes were chosen because they are common outcomes and represented outcomes that were not primarily of infectious etiology and therefore less likely to be associated with the vaccination. We again used a CoxPH model including vaccination as a time-dependent variable, stratified by year of birth adjusting for the same confounders and for the correlation between patients of the same practice. Furthermore, we compared the underlying causes of death for those who died during the follow-up period.

### 2.4. Governance

Use of anonymized linked data from SAIL was granted under Information Governance Review Panel (IGRP) 0938. All data management and analysis were performed on the secure research platform from SAIL.

## 3. Results

The study population comprised 336,341 people (total of 2,284,603 person-years of follow-up), of whom 161,428 (48%) were born after 1943 and 162,142 (48%) were male. During follow-up, 155,972 (46%) were exposed to the vaccine, 53,822 (16%) died, and 18,570 (5.5%) were diagnosed with dementia. People less likely to be vaccinated were female; had been diagnosed with rheumatic disease, perivascular disease, MI, liver disease, cerebrovascular disorders, or coronary heart disease, were living in a care home, or were frail. Renal disease, diabetes, and chronic pulmonary disease, prior vaccination, and higher socioeconomic status were positively associated with shingles vaccination (Table 1).

Most people vaccinated between 2013 and 2019 were vaccinated at age 70 years; a further group of people were vaccinated between age 77 and 79. Especially over the later years, vaccinations at ages other than the suggested 70 and 79 were more common (Supplementary Figure S1). Accordingly, the vaccination rate differed widely between those born before and after 1943.

Compared to people not vaccinated during follow-up, people who were exposed to the shingles vaccine were at lower risk of being diagnosed with dementia (adjusted hazard ratio, aHR: 0.72; 95% CI: 0.69 to 0.75; univariable HR: 0.78; 95% CI: 0.75 to 0.81) (Figure 1A and Table S2).

**Figure 1.**
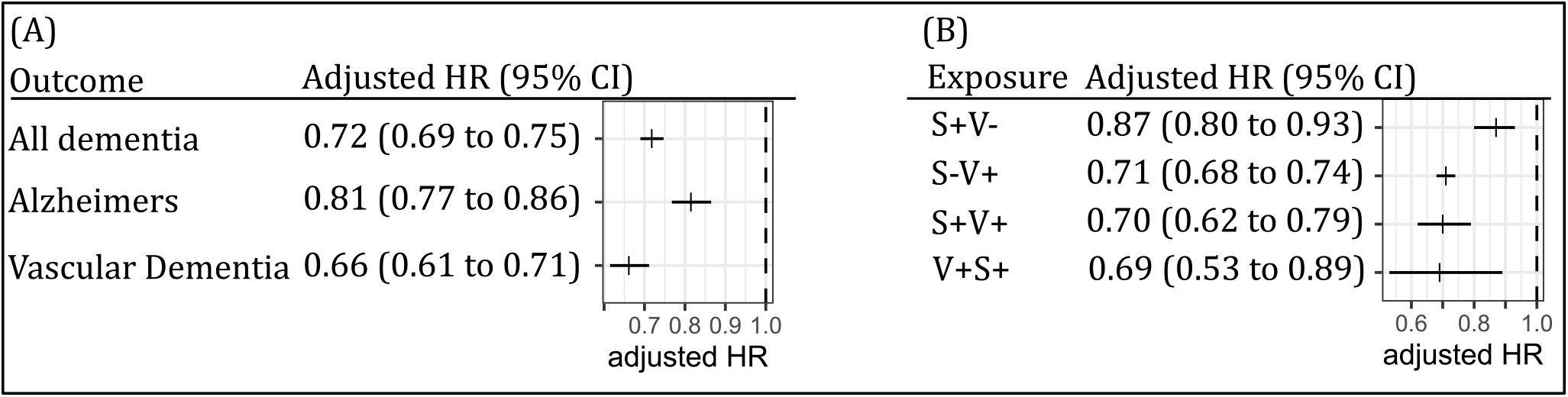
Results (adjusted HR and 95% CI) from the multivariable Cox proportional hazard model of the association between exposure to shingles vaccination and dementia. (A) Classified by type of dementia (Alzheimer’s disease; and vascular dementia). The comparison group (hazard ratio, HR = 1.0) was not vaccinated. (B) Classified by exposure (S+V-: shingles, not vaccinated; S-V+: no shingles, vaccinated; S+V+: first shingles, then vaccinated; V+S+: first vaccinated, then shingles). The comparison group is no shingles, not vaccinated. Full results of both models are given in the supplementary data online.

That association varied slightly with the two dementia subtypes: People exposed to the shingles vaccine were at lower risk of being diagnosed with AD (aHR: 0.81; 95% CI: 0.77 to 0.86) and at lower risk of being diagnosed with VaD (aHR: 0.66; 95% CI: 0.61 to 0.71) (Figure 1A, and Tables S3 and S4). Shingles vaccination was associated with a substantial reduction in subsequent shingles diagnosis (aHR: 0.43; 95% CI: 0.41 to 0.45) (Figure 2); however, we could not find any evidence that the association of shingles vaccination with dementia was mediated by a reduction in shingles diagnosis. Compared to people not vaccinated with no shingles diagnosis, those vaccinated with no shingles diagnosis had 0.71 times lower hazard (aHR: 0.71; 95% CI: 0.68 to 0.74), whereas those vaccinated with a subsequent diagnosis of shingles (’vaccine failures’) had a similar, 0.69 times lower hazard (aHR: 0.69; 95% CI: 0.53 to 0.89) (Figure 1B and Table S5).

**Figure 2.**
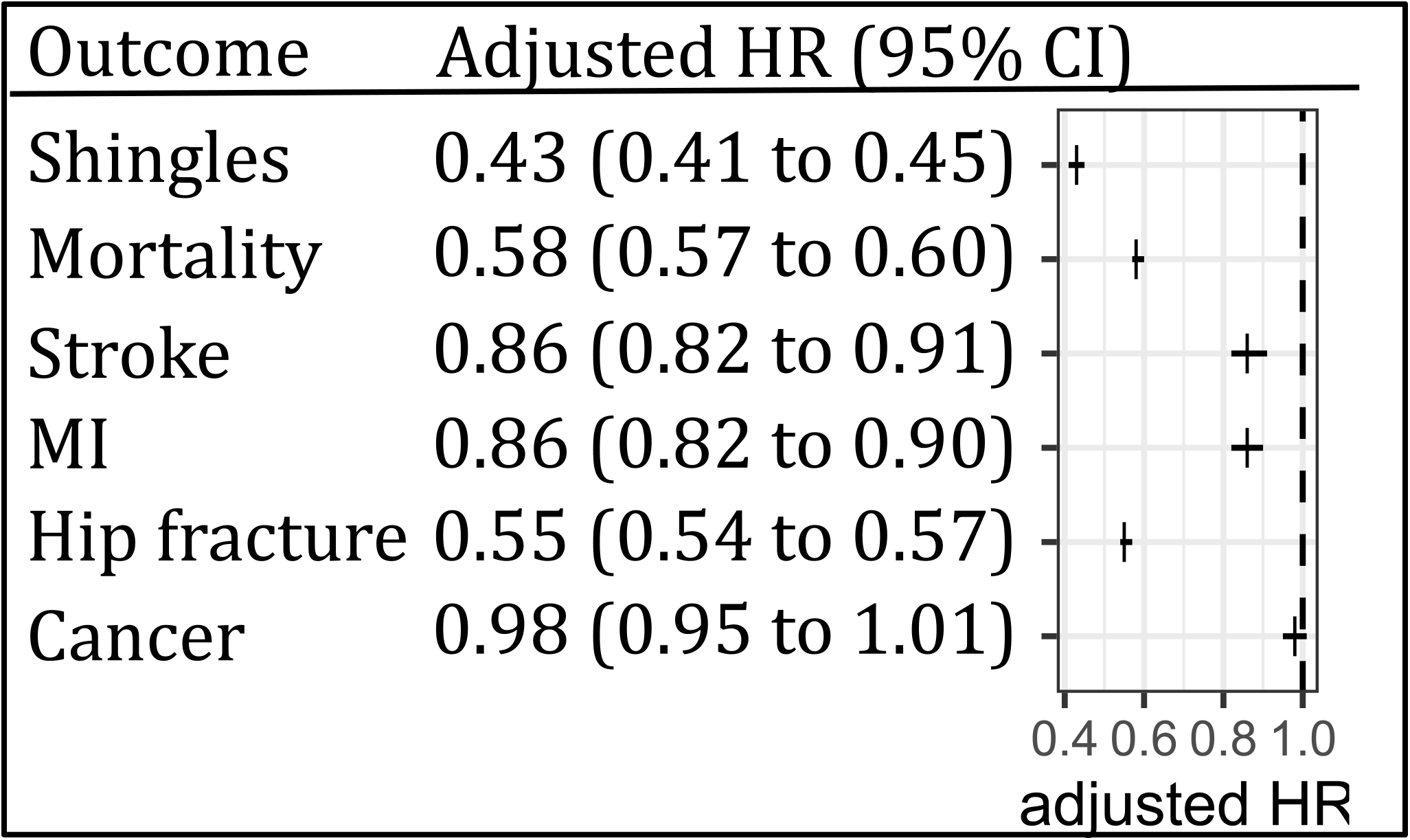
Results (adjusted hazard ratio, HR; and 95% CI) from the multivariable Cox proportional hazard models of the association between exposure to shingles vaccination and shingles, all-cause mortality, stroke, myocardial infarction, hip fracture, and cancer. The comparison group (HR = 1.0) was not vaccinated.

Compared to people not vaccinated during follow-up, people who were exposed to the shingles vaccine were at lower risk of being diagnosed with MI (aHR: 0.86; 95% CI 0.82 to 0.90), stroke (aHR: 0.86; 95% CI 0.82 to 0.91), or hip fracture (aHR: 0.55; 95% CI 0.54 to 0.57; there was no difference in the risk of being newly diagnosed with cancer during follow-up (aHR: 0.98; 95% CI 0.95 to 1.01). Similarly, people who were exposed to the shingles vaccine were at lower risk of all-cause mortality (aHR: 0.58; 95% CI 0.57 to 0.60) (Figure 2). A purely descriptive/exploratory analysis of the underlying causes for mortality in those who died during follow-up did not reveal differences between vaccinated and non-vaccinated individuals (Figure S2).

## 4. Discussion

We analyzed the association of shingles vaccination with incident dementia in those vaccinated in Wales between 2013 and 2020 in an observational cohort study using retrospectively collected national health data. People exposed to the vaccine had a 39% reduced hazard of dementia diagnosis following vaccination. This association is similar to the study by Scherrer *et al*.[3] who found a 43% reduction in dementia in those vaccinated against shingles. It compares favorably to the modifiable risk factors for dementia in later life summarized in Livington[19]: smoking (relative risk, RR: 1.6), depression (RR: 1.9), social isolation (RR: 1.6), physical inactivity (RR: 1.4), diabetes (RR: 1.5), and air pollution (RR: 1.1).

The reduction in dementia in those exposed to the vaccine was slightly more pronounced for VaD than for AD. This is an interesting finding because, to our knowledge, none of the vaccine studies have separately analyzed dementia subtypes. Although the risk of misclassification of dementia subtypes in RCHR is high[15], we have no evidence for differential misclassification (misclassification of dementia subtype is independent of vaccine status). If true, our findings point towards an association between shingles vaccination and cerebrovascular pathologies[20,21] rather than an association of vaccination with the pathological accumulation of toxic proteins in the brain such as amyloid-beta peptide and tau. Further evidence for an association of vaccination with cerebrovascular health was our finding that shingles vaccination was additionally associated with a reduction of stroke incidence.

Shingles vaccination was associated with a substantial reduction in shingles incidence (aHR: 0.43), which is slightly better than the vaccine effect reported by Blom *et al*. [22] (aHR = 0.66, 95% CI: 0.55-0.78). However, the reduced hazard of dementia in those vaccinated against shingles was probably not mediated by a reduction in shingles incidence because the HR in people not vaccinated without shingles was similar to the HR in those vaccinated without shingles, as well as the HR in those vaccinated without subsequent shingles (’vaccine failures’). This result points to a non-specific effect of shingles vaccination on dementia incidence rather than a direct effect via shingles reduction. However, our results must be interpreted carefully because the total follow-up time of people who were vaccinated and had a subsequent diagnosis of shingles was low, causing wide confidence intervals in the estimate.

People exposed to the shingles vaccine were at lower risk of all-cause mortality (aHR: 0.58), MI, (aHR: 0.86), stroke (aHR: 0.86), and hip fracture (aHR: 0.55) – but not cancer (aHR: 0.98) – and the aHRs were similar in size to the aHR for the association of vaccination with dementia. This result could indicate a non-specific effect of the shingles vaccination[23,24]. We have carefully considered the possibility of a potential mechanism that explains our findings, particularly in AD. There has been escalating interest in the possibility that AD is triggered by infection[25] and that the signature protein of AD brain, Aβ peptide, has antimicrobial activity, and thus may be a consequence rather than a cause of AD[26]. One potential interpretation of our results is therefore that the live attenuated VZV vaccine acts as an adjuvant that plays a role in the immune responses against microbes. This interpretation is supported by (i) documented immune cross-protection where infection with one pathogen can alleviate the disease caused by a second unrelated pathogen[24], and (ii) precedent: an immune adjuvant alone (alum) was reported to retard AD development[27], and a potent adjuvant vaccine (killed attenuated *Mycobacterium*, BCG) was reported to reduce AD rates in bladder cancer patients[2]. The adjuvant theory should be considered carefully, alongside other theories, as a potential explanation for the negative association between VZV vaccination and incident dementia. However, the additional outcomes we have chosen (cancer, MI, stroke, hip fracture, and all-cause mortality) are primarily not driven by infectious pathogens, and differences in mortality were not more pronounced in diseases with a predominantly infectious etiology (e.g., respiratory diseases). Furthermore, the association we observed was stronger for VaD compared to AD. Therefore, our results might indicate a selection bias, with people getting vaccinated having a higher (healthy) life expectancy at the time of vaccination, and thus less likely to be at risk of imminent diagnosis of dementia. Indeed, non-specific vaccine effects such as lower mortality have previously been described in observational cohort studies of vaccine efficacy by Simonsen *et al*.[5], who attributed the association to a ’frailty selection bias’. To control for frailty selection bias we adjusted for frailty (EFI) between age 65 and 70, for care home residency, and for multiple diseases that make up the Charlson comorbidity index. All these variables were associated with lower rates of vaccination (Table 1) and increased rates of dementia (Tables S2–S4); however, adjusting for them in the dementia model (or any of the other outcomes) did not drastically modify the estimates.

Selection bias may be the main limitation of our study. Similarly to other observational cohort studies, the reason why some people were exposed to the vaccine, whereas others were not, remains unknown. Although we controlled for ’frailty’, we cannot exclude with confidence that people not being vaccinated might have a lower healthy life expectancy. This observation would be supported by results from vaccine efficacy studies for Zostavax, which did not show any significant difference in mortality between those exposed to the vaccine and those exposed to a placebo[28]. Furthermore, even though our study population was large and representative of the Welsh population, the average follow-up period was rather short. This was due to the introduction of the vaccine in 2013, which gave us a maximum follow-up time of approximately 6 years (until age 76).

## 5. Conclusions

Our study shows a clear association between shingles vaccination and reduced dementia incidence, which is consistent with other observational cohort studies. As such, non-exposure to a vaccine, be it a shingles vaccine or Tdap, could potentially be used as an early warning sign of deteriorating health, and it would be interesting to analyze whether vaccine exposure could be used in predictive modeling. Although we cannot exclude a true effect, our results lead us to suspect that the association reflects frailty selection bias rather than a genuine vaccine effect. The unexpected greater association with VaD than with AD, the unexpected result that a diagnosis of shingles was not a mediating variable, the non-specific effects of vaccination on other diseases and mortality and the fact that other vaccines such as Tdap and BCG were also associated with reduced dementia in other studies points more to a selection bias associated with vaccination than a genuine vaccine effect. We further encourage consideration of and research into potential theories that may explain the negative association with incident dementia and shed light on whether this may reflect a genuine vaccine effect.

## Data Availability

All data used for analysis is accessible on the SAIL databank under governanace from Information Governance Review Panel

## Acknowledgments

This work was supported by the Benter Foundation.

## Supplementary material online

Supplementary material associated with this article can be found, in the online version, at DOI XXX-XXX-XXX

