## Supplemental material for "Reduced dementia incidence following varicella zoster vaccination in Wales 2013–2020"

**Supplementary Material Online**

1. Counts of vaccinations per year by age group (Figure 1)
2. Code lists for classification (dementia and vaccination, only)(Table 1)
3. Detailed results from the multivariable Cox proportional hazard models
   a. All dementia (Table 2)
   b. AD (Table 3)
   c. VaD (Table 4)
   d. By shingles diagnosis (Table 5)
4. Proportion of mortality causes by ICD code (Figure 2)

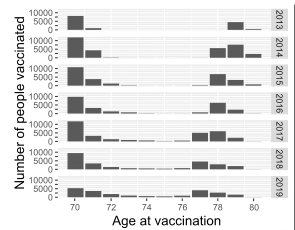

**Figure 1.** Number of people in the study population vaccinated with a shingles vaccine by age and year of vaccination between 2013 and 2019.

**Table 1.** Codes used for classification of dementia and shingles vaccination in data from primary and secondary care. We used notation from regular expression, where ^1461 denotes all codes starting with 1461; ^3AE[3–6] denotes all codes starting with 3AE followed by any number between 3 and 6; and ^F110[.01] denotes all codes starting with F110 followed by either a dot, 0, or 1.

|  | Dementia | Shingles vaccination |
| --- | --- | --- |
| Primary care^*^ | ^1461; ^38C13; ^3AE[3-6]; ^66h; ^6AB; ^8BM02; ^8CMG2; ^8CMZ[.0-3]; ^8CSA.; ^8Hla; ^8Iae2; ^9hD[.01]; ^9Ou; ^A411[.0]; ^E00[.0-4]; ^E012; ^E02y1; ^E041; ^Eu00[.012z]; ^Eu01[.0-3yz]; ^Eu02[.0-5yz]; ^Eu041; ^Eu10[67]; ^F110[.01]; ^F11[1268]; ^F11x[279]; ^F11y2; ^F21y2; ^ Fyu30 | ^65FY. |
| Hospital admission^†^ | ^F0[0-3]; ^F051; ^G30; ^G31[018]; ^I673 |  |
| Prescription^*^ |  | ^n4v.. |

^*^READ version 2 codes

^†^ICD 10 codes

**Table 2**. Results (adjusted hazard ratio and 95% CI) from the multivariable Cox proportional hazard model^*^ on the association of shingles vaccination with dementia.

| Variable | Level | aHR (95% CI; *p*-value) |
| --- | --- | --- |
| Vaccinated | No |  |
|  | Yes | 0.72 (0.69 to 0.75); p<0.01 |
| Sex | Male |  |
|  | Female | 0.97 (0.94 to 1); p=0.04 |
| WIMD^†^ | 1 |  |
|  | 2 | 0.93 (0.89 to 0.98); p<0.01 |
|  | 3 | 0.87 (0.83 to 0.91); p<0.01 |
|  | 4 | 0.81 (0.77 to 0.85); p<0.01 |
|  | 5 | 0.74 (0.7 to 0.77); p<0.01 |
| Care home | No |  |
|  | Yes | 4.1 (3.27 to 5.13); p<0.01 |
| Frailty^‡^ |  | 1.59 (1.55 to 1.62); p<0.01 |
| Prior vaccination | No |  |
|  | Yes | 1.02 (0.98 to 1.06); p=0.39 |
| Diabetes | No |  |
|  | Yes | 1.04 (1 to 1.09); p=0.06 |
| Cancer | No |  |
|  | Yes | 0.94 (0.88 to 1); p=0.04 |
| Cerebro-vascular disease | No |  |
|  | Yes | 1.53 (1.43 to 1.62); p<0.01 |
| COPD | No |  |
|  | Yes | 0.87 (0.83 to 0.91); p<0.01 |
| Chronic heart disease | No |  |
|  | Yes | 1.02 (0.92 to 1.14); p=0.66 |
| Chronic liver disease | No |  |
|  | Yes | 1.26 (0.97 to 1.63); p=0.08 |
| Myocardial infarctation | No |  |
|  | Yes | 0.93 (0.84 to 1.03); p=0.16 |
| Peptic ulcer | No |  |
|  | Yes | 1.09 (0.94 to 1.25); p=0.25 |
| Perivascular disease | No |  |
|  | Yes | 1.16 (1.05 to 1.28); p<0.01 |
| Renal disease | No |  |
|  | Yes | 0.86 (0.81 to 0.91); p<0.01 |
| Rheumatic disease | No |  |
|  | Yes | 0.98 (0.89 to 1.08); p=0.65 |

^*^The model was stratified by year of birth (<1943 and ≥1943) and additionally adjusted for correlation between patients of the same GP practice (included as random effect).

^†^Welsh Index of multiple deprivation (quintiles); 1 is most deprived.

^‡^Included as continuous variable as (EFI + 0.1)/0.1 (best fitting fractional polynomial)

**Table 3.** Results (adjusted hazard ratio and 95% CI) from the multivariable Cox proportional hazard model^*^ on the association of shingles vaccination with Alzheimer’s dementia.

| **Variable** | **Level** | **Adjusted HR (95% CI)** |
| --- | --- | --- |
| Vaccinated | No |  |
|  | Yes | 0.81 (0.77 to 0.86); p<0.01 |
| Sex | Male |  |
|  | Female | 1.18 (1.13 to 1.23); p<0.01 |
| WIMD^†^ | 1 |  |
|  | 2 | 0.95 (0.88 to 1.02); p=0.13 |
|  | 3 | 0.85 (0.79 to 0.92); p<0.01 |
|  | 4 | 0.8 (0.74 to 0.86); p<0.01 |
|  | 5 | 0.81 (0.75 to 0.87); p<0.01 |
| Care home | No |  |
|  | Yes | 1.7 (0.96 to 2.99); p=0.07 |
| Frailty^‡^ |  | 1.42 (1.37 to 1.47); p<0.01 |
| Prior vaccination | No |  |
|  | Yes | 1.17 (1.1 to 1.24); p<0.01 |
| Diabetes | No |  |
|  | Yes | 0.98 (0.91 to 1.04); p=0.46 |
| Cancer | No |  |
|  | Yes | 0.97 (0.88 to 1.05); p=0.44 |
| Cerebro-vascular disease | No |  |
|  | Yes | 0.9 (0.79 to 1.01); p=0.08 |
| Chronic obstructive pulmonary disease | No |  |
|  | Yes | 0.91 (0.85 to 0.98); p=0.01 |
| Chronic heart disease | No |  |
|  | Yes | 0.75 (0.62 to 0.92); p=0.62 |
| Chronic liver disease | No |  |
|  | Yes | 0.89 (0.55 to 1.43); p=0.99 |
| Myocardial infarctation | No |  |
|  | Yes | 0.86 (0.73 to 1.02); p<0.01 |
| Peptic ulcer | No |  |
|  | Yes | 1 (0.79 to 1.26); p<0.01 |
| Perivascular disease | No |  |
|  | Yes | 0.83 (0.69 to 1); p<0.01 |
| Renal disease | No |  |
|  | Yes | 0.85 (0.77 to 0.93); p<0.01 |
| Rheumatic disease | No |  |
|  | Yes | 0.82 (0.7 to 0.96); p<0.01 |

^*^The model was stratified by year of birth (<1943 and >=1943) and additionally adjusted for correlation between patients of the same GP practice (included as random effect).

^†^Welsh Index of multiple deprivation (quintiles); 1 is most deprived.

^‡^Included as continuous variable as (EFI + 0.1)/0.1 (best-fitting fractional polynomial).

**Table 4.** Results (adjusted hazard ratio, HR; and 95% CI) from the multivariable Cox proportional hazard model^*^ on the association between shingles vaccination and vascular dementia

| Variable | Level | Adjusted HR (95% CI) |
| --- | --- | --- |
| Vaccinated | No |  |
|  | Yes | 0.66 (0.61 to 0.71); p<0.01 |
| Sex | Male |  |
|  | Female | 0.8 (0.76 to 0.84); p<0.01 |
| WIMD^†^ | 1 |  |
|  | 2 | 0.91 (0.84 to 0.99); p=0.02 |
|  | 3 | 0.82 (0.76 to 0.89); p<0.01 |
|  | 4 | 0.76 (0.7 to 0.83); p<0.01 |
|  | 5 | 0.65 (0.59 to 0.71); p<0.01 |
| Care home | No |  |
|  | Yes | 1.63 (0.94 to 2.82); p=0.08 |
| Frailty^‡^ |  | 1.75 (1.68 to 1.82); p<0.01 |
| Prior vaccination | No |  |
|  | Yes | 0.97 (0.91 to 1.04); p=0.47 |
| Diabetes | No |  |
|  | Yes | 1.2 (1.12 to 1.28); p=0.15 |
| Cancer | No |  |
|  | Yes | 0.93 (0.84 to 1.03); p<0.01 |
| Cerebrovascular disease | No |  |
|  | Yes | 2.47 (2.27 to 2.7); p=0.3 |
| Chronic obstructive pulmonary disease | No |  |
|  | Yes | 0.87 (0.8 to 0.94); p=0.08 |
| Chronic heart disease | No |  |
|  | Yes | 1.19 (1.02 to 1.4); p<0.01 |
| Chronic liver disease | No |  |
|  | Yes | 1.26 (0.81 to 1.96); p<0.01 |
| Myocardial infarctation | No |  |
|  | Yes | 1.06 (0.91 to 1.23); p<0.01 |
| Peptic ulcer | No |  |
|  | Yes | 1.22 (0.98 to 1.51); p<0.01 |
| Perivascular disease | No |  |
|  | Yes | 1.48 (1.29 to 1.71); p<0.01 |
| Renal disease | No |  |
|  | Yes | 0.85 (0.77 to 0.93); p<0.01 |
| Rheumatic disease | No |  |
|  | Yes | 0.93 (0.79 to 1.1); p<0.01 |

^*^The model was stratified by year of birth (<1943 and >=1943) and additionally adjusted for correlation between patients of the same GP practice (included as random effect).

^†^Welsh Index of multiple deprivation (quintiles); 1 is most deprived.

^‡^Included as continuous variable as (EFI + 0.1)/0.1 (best fitting fractional polynomial)

Figure 2. Underlying cause (ICD10 sections) of death by vaccine status. ICD10 sections: A, Certain infectious and parasitic diseases; C, malignant neoplasms; D, malign neoplasms and diseases of the blood and blood-forming organs and certain disorders involving the immune mechanism; E, endocrine, nutritional, and metabolic diseases; F, mental and behavioural disorders; G, diseases of the nervous system; I, diseases of the circulatory system; J, diseases of the respiratory system; K, diseases of the digestive system; L, diseases of the skin and subcutaneous tissue; M, diseases of the musculoskeletal system and connective tissue; N, diseases of the genitourinary system; V–Y, external causes of morbidity and mortality. For reasons of data protection, underlying causes B, H, O–U and Z have been grouped into group Z (any other) (source: WHO ICD10 browser).

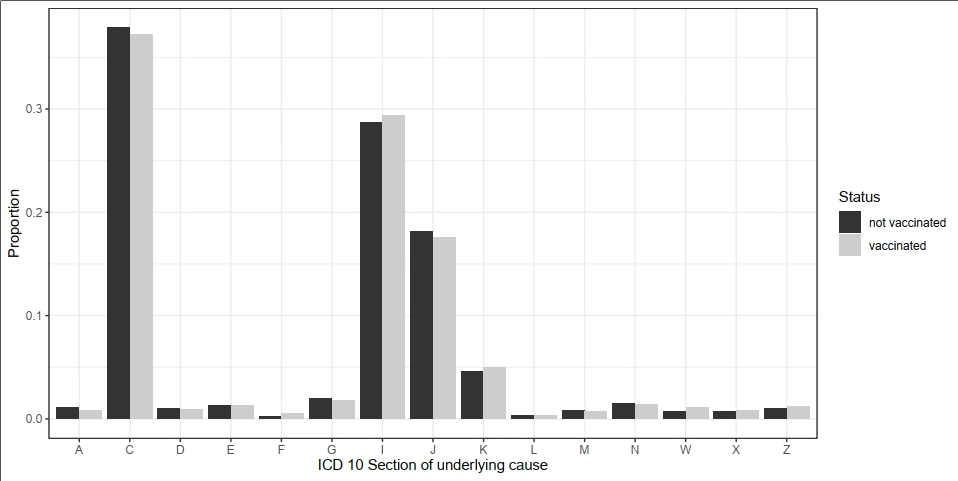
